# Association Between Birth Region and Time to Tuberculosis Diagnosis After US Entry Among Non–US-Born Persons

**DOI:** 10.1101/2020.08.02.20160135

**Authors:** Amish Talwar, Rongxia Li, Adam J. Langer

**Affiliations:** Centers for Disease Control and Prevention, Atlanta, Georgia, USA

**Author notes:** Address for correspondence: Amish Talwar, Centers for Disease Control and Prevention, 1600 Clifton Rd NE, Mailstop US12-4, Atlanta, GA, 30329, USA.

## Abstract

Approximately 90% of US tuberculosis (TB) cases among non–US-born persons are attributable to progression of latent TB infection to TB disease. Using survival analysis, we investigated if birthplace is associated with time of progression to TB disease among non–US-born persons. We derived a Cox regression model comparing differences in time to TB diagnosis after US entry among 19 global birth regions, adjusting for sex, birth year, and age at diagnosis. Compared with persons from Western Europe, the adjusted hazard rate of developing TB was significantly higher (p ≤0.05) for persons from all other regions, except North America and Northern Europe, and highest among persons from Middle Africa (adjusted hazard ratio = 7.0; 95% confidence interval: 6.5–7.4). Time to TB diagnosis among non–US-born persons therefore varied by birth region, which represents an important prognostic indicator for progression to TB disease.

## Introduction

The majority of incident tuberculosis (TB) cases in the United States occurs among non– US-born persons. During 2018, 70.2% of TB disease cases occurred among non–US-born persons, and 46.6% of those cases were diagnosed ≥10 years after arrival to the United States (*1*). Although TB disease can result from recent person-to-person transmission, it is more commonly the result of progression of latent TB infection (LTBI) to TB disease. LTBI is a form of TB in which a person is infected with *Mycobacterium tuberculosis*, the causative agent of TB, but remains asymptomatic and noncontagious (*2*); if left untreated, LTBI can progress to TB disease among ≤10% of persons with LTBI within their lifetime (*2*). Because >85% of TB disease cases among non–US-born persons residing in the United States are attributed to progression of LTBI to TB disease (*3,4*), a substantial proportion of TB cases among non–US-born persons is likely the result of progression of LTBI acquired before US arrival.

Consequently, the Centers for Disease Control and Prevention (CDC) recommend that efforts to eliminate TB in the United States, in part, focus on LTBI detection and treatment among non– US-born persons (*5*).

Understanding the factors associated with LTBI progression can help guide these efforts by identifying persons with LTBI who are at greatest risk of developing TB disease, allowing TB prevention resources to be concentrated on the populations at highest risk. Although persons recently infected with TB and persons with weakened immune systems are at higher risk for progressing to TB disease (*6*), the factors affecting the time required to develop TB disease remain unclear, particularly for non–US-born persons. Knowing such information can help public health officials target interventions for LTBI testing and treatment for these persons before LTBI progresses to TB disease. One study found the risk of developing TB disease among non–US-born persons decreased with increasing time since US entry (*7*). However, non–US-born persons are a heterogenous population who differ in health status based on country of origin (*8*), and the effect of birth country on progression from LTBI to TB disease remains unclear. To answer this question, we compared the time necessary for non–US-born persons to develop TB disease after arrival in the United States according to birthplace.

## Methods

Using US national TB surveillance data for 2011–2018, we assessed time to TB disease diagnosis among non–US-born persons after US entry according to geographical birth region (see “Research Design and Variables”); we categorized birth countries into regions because of the high number of countries represented among non–US-born persons with TB disease. We assessed time to TB disease diagnosis as time from US entry to the time the TB case was reported to a local or state health department, and we excluded persons with TB disease attributed to recent transmission (*3*) to focus on persons whose TB disease was most likely caused by progression of LTBI acquired in their birth countries. We compared these times by using bivariate and multivariate survival analysis, and we adjusted for age at TB disease diagnosis, sex, and birth year.

### Study Population

We derived the study population for this analysis from CDC’s National Tuberculosis Surveillance System (NTSS), which has been collecting information on TB disease cases in the United States since 1953 (*9*). Case reports include demographic, clinical, and risk factor data. NTSS’s most recent TB surveillance case definition as of 2009 is available in the 2018 US TB surveillance report (*1*). For this study, we examined cases reported to NTSS from January 2011 through December 2018 among non–US-born persons (i.e., persons born outside the United States or its territories for whom neither parent was a US citizen) (birth countries and territories are listed in Appendix A). We excluded cases reported from the US territories or states freely associated with the United States (Figure 1).

**Figure 1.**
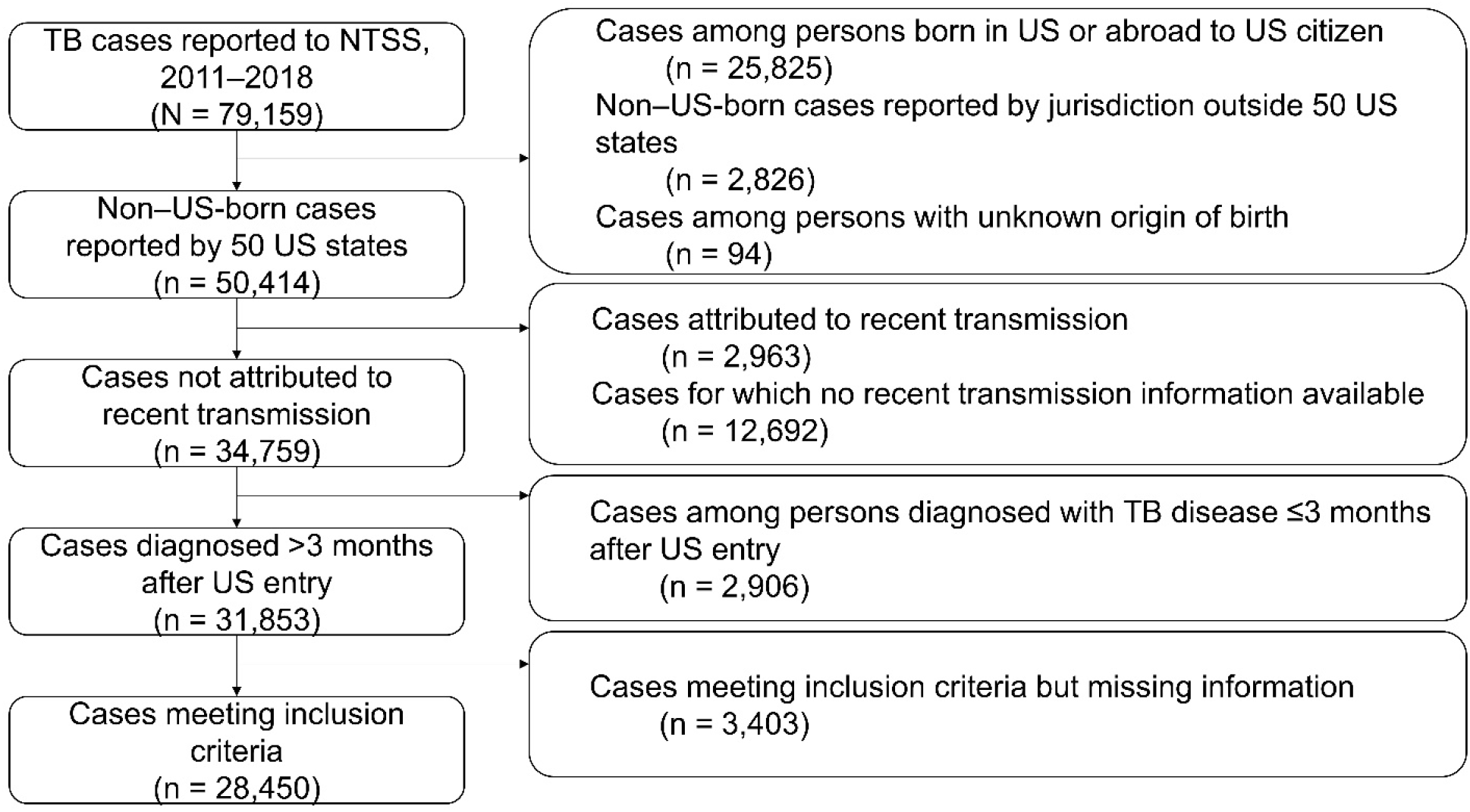
Flow chart of analysis cohort selection process.

We also excluded cases attributed to recent TB transmission in the United States by using a previously published method that uses NTSS and genotypic data to determine likelihood of recent transmission. This plausible-source case method attributes cases to recent transmission if at least one plausible source case for the case of interest is identified in NTSS (*3*). With this method, cases are either attributed to recent transmission, not attributed to recent transmission, or receive neither designation if they lack the necessary information to assess for recent transmission (e.g., missing genotype data). By excluding cases attributed to recent transmission, we were able to analyze cases that were more likely the result of LTBI progression. We further excluded cases not attributed to recent transmission among persons for whom time from arrival to TB disease diagnosis was ≤3 months because TB disease among such persons can represent disease that was present at time of US entry rather than LTBI reactivation after US arrival; excluding these cases also helps account for variability in TB disease screening overseas before US entry.

Finally, we excluded cases with missing data and observations with nonsensical values for a category (i.e., months to TB disease diagnosis <0, non–US-born persons with “United States” listed as birth country, or age less than number of months to TB disease diagnosis).

### Research Design and Variables

We performed a bivariate analysis and a multivariate analysis examining the association between birth country and time from initial US arrival to TB disease diagnosis, which was our main outcome variable. In addition to birth country, we examined the following NTSS case demographic variables as additional covariates for our analyses: age at TB disease diagnosis, sex, and birth year. We performed the bivariate analysis to assess the association between time to TB disease diagnosis and those variables individually, and we performed the multivariate analysis to account for the effects of the additional demographic variables on the association between birth country and time to TB disease diagnosis. For our analyses, time to TB disease diagnosis was the number of months spent in the United States before TB disease diagnosis, which we derived by subtracting the date of initial US entry from the date that the TB disease case was reported. We used the case report date because information regarding actual disease diagnosis date was unavailable through NTSS; the report date represents the earliest notification to a local public health agency that the patient might have TB disease. Because >200 non–US countries and territories of birth were reported to NTSS during the study timeframe, we used the United Nations M49 standard, which categorizes countries and territories according to geographic location and level of development, to divide these countries into 19 regions for ease of statistical analysis (*10,11*) (see “Birth Country Categorization” in Appendix B). For our bivariate analysis, we categorized age into 6 categories, but we kept age as a continuous variable for our multivariate analysis. We derived birth year by subtracting age from the year the case was reported, and we categorized birth year into 6 categories; we included birth year in our analysis to adjust for a previously observed birth year cohort effect among non–US-born persons with TB disease reported to NTSS (*12*). Finally, we excluded race/ethnicity as a covariate because of multicollinearity with birth region (see “Exclusion of race/ethnicity variable” in Appendix B).

### Statistical Analysis

We began by determining the number of patients who had TB disease not attributed to recent transmission in each demographic category and determined the median number of months (with interquartile range [IQR]) that these patients spent in the United States before receiving a TB disease diagnosis. We also determined the median number of months according to birth region and median age. We constructed a crude bivariate Kaplan-Meier survival curve to visually demonstrate the overall distribution of time to TB diagnosis since US entry. We then constructed bivariate Kaplan-Meier curves stratified by each demographic covariate. We tested for statistically significant differences between curves at p ≤0.05 by using the log rank test and reported global p values.

We then used Cox regression to examine the association of time to TB disease diagnosis and birth region, adjusting for the effects of age at diagnosis, sex, and birth year. We tested the proportional hazards assumption (see “Cox regression assumptions” in Appendix B), and because our data are likely double-truncated, we applied a correction for double-truncation to our sample set (*13*) (see “Double-truncation” in Appendix B). We reported adjusted hazard ratios (aHRs) with associated 95% confidence intervals (CIs) and p values from our Cox regression analysis.

Because the time after US entry at which persons who enter the United States with TB disease typically receive a diagnosis is unclear, previous studies have used a range of post-entry times to define cases as disease missed on entry as opposed to LTBI reactivation (*14*–*16*).

Therefore, we performed a sensitivity analysis to assess the effect of extending the exclusion window from 3 months to 6 months for TB disease missed at time of entry for our Cox regression analysis. Additionally, because TB in a child can be considered a sentinel event that indicates recent transmission (*17*), we performed a sensitivity analysis to assess the effect of excluding children aged <5 years on the basis of previous research demonstrating that recent transmission is most commonly observed among children in that age group (*3,4*).

We performed all statistical analyses using R version 3.6.1, and we used the R code developed by Rennert and Xie to correct for double truncation (*13*). All data were collected as part of routine disease surveillance and were not part of human subjects research requiring institutional review board approval.

## Results

During 2011–2018, 79,159 TB cases were reported to NTSS (Figure 1). Of those cases, 28,745 cases were among US-born persons, were not reported from one of the 50 states or the District of Columbia, or did not have a known origin of birth. We also excluded 15,655 cases that were either attributed to recent transmission or had no information regarding recent transmission available; we excluded 2,906 cases for which the time to TB diagnosis was ≤3 months. Finally, we excluded 3,403 cases with missing information for ≥1 of the variables considered; because only approximately 10% of the included cases had missing data for ≥1 variable, we performed listwise deletion of those cases for our analyses. As a result, we included 28,450 cases for our analyses.

The median number of months that a non–US-born person spent in the United States before receiving a diagnosis of TB disease not attributed to recent transmission was 144 months (IQR: 52–294 months). Persons from Middle Africa had the lowest median number of months until TB diagnosis (26 months), and persons from Western Europe had the highest median number of months (541 months) (Table 1). The median patient age was 48 years (IQR: 32–65 years). The highest proportion of non–US-born persons who developed TB disease not attributed to recent transmission after arrival to the United States emigrated from Asia (Table 2); the subregion or intermediary region with the highest proportion of non–US-born persons who received a TB diagnosis in the United States not attributed to recent transmission was South-eastern Asia.

**Table 1.** Median time to diagnosis of tuberculosis disease not attributed to recent transmission for non–US-born persons by region — United States, 2011–2018

| **Region** | **Median time to tuberculosis disease diagnosis (months)** |
| --- | --- |
| *Africa* | 53 |
| Eastern Africa | 62 |
| Middle Africa | 26 |
| Northern Africa | 64 |
| Southern Africa | 77 |
| Western Africa | 46 |
| *Americas* | 162 |
| Caribbean | 127 |
| Central America | 175 |
| Northern America* | 353 |
| South America | 142 |
| *Asia* | 157 |
| Central Asia | 60 |
| Eastern Asia | 211 |
| South-eastern Asia | 190 |
| Southern Asia | 76 |
| Western Asia | 83 |
| *Europe* | 214 |
| Eastern Europe | 178 |
| Northern Europe | 455 |
| Southern Europe | 223 |
| Western Europe | 541 |
| *Oceania* | 67 |

\* Excludes the United States of America.

**Table 2.** Adjusted hazard ratio (aHR) estimates of tuberculosis disease diagnosis for non–U.S-born persons with tuberculosis disease not attributed to recent transmission, adjusting for age at diagnosis, sex, and birth year (n = 28,450) — United States, 2011–2018

| Region | No. (%) | aHR | 95% CI | p value |
| --- | --- | --- | --- | --- |
| *Africa* | 2,910 (10.2) |  |  |  |
| Eastern Africa | 1,627 (5.7) | 3.91 | (3.67–4.17) | <0.01 |
| Middle Africa | 286 (1.0) | 7.02 | (6.53–7.56) | <0.01 |
| Northern Africa | 126 (0.4) | 3.43 | (3.15–3.75) | <0.01 |
| Southern Africa | 56 (0.2) | 4.19 | (3.74–4.70) | <0.01 |
| Western Africa | 815 (2.9) | 4.84 | (4.54–5.17) | <0.01 |
| *Americas* | 9,719 (34.2) |  |  |  |
| Caribbean | 1,241 (4.4) | 2.82 | (2.65–3.01) | <0.01 |
| Central America | 7,114 (25.0) | 2.03 | (1.91–2.15) | <0.01 |
| Northern America* | 20 (0.1) | 1.13 | (0.99–1.29) | 0.07 |
| South America | 1,344 (4.7) | 2.85 | (2.68–3.03) | <0.01 |
| *Asia* | 14,999 (52.7) |  |  |  |
| Central Asia | 45 (0.2) | 4.71 | (4.17–5.32) | <0.01 |
| Eastern Asia | 2,924 (10.3) | 2.88 | (2.71–3.07) | <0.01 |
| South-eastern Asia | 7,812 (27.5) | 3.06 | (2.89–3.26) | <0.01 |
| Southern Asia | 4,026 (14.2) | 4.29 | (4.04–4.57) | <0.01 |
| Western Asia | 192 (0.7) | 3.34 | (3.09–3.62) | <0.01 |
| *Europe* | 669 (2.4) |  |  |  |
| Eastern Europe | 328 (1.2) | 2.77 | (2.58–2.98) | <0.01 |
| Northern Europe | 47 (0.2) | 1.12 | (1.01–1.23) | 0.03 |
| Southern Europe | 226 (0.8) | 1.25 | (1.17–1.35) | <0.01 |
| Western Europe† | 68 (0.2) | NA | NA | NA |
| *Oceania* | 153 (0.5) | 2.78 | (2.56–3.02) | <0.01 |
| Australia and New Zealand | 1 (0.004) | NA | NA | NA |
| Melanesia | 10 (0.04) | NA | NA | NA |
| Micronesia | 113 (0.4) | NA | NA | NA |
| Polynesia | 29 (0.1) | NA | NA | NA |

Abbreviations: CI: confidence interval; NA: not available.
\* Excludes the United States of America.
† Indicates reference value.

Figures 2 and 3 display the Kaplan-Meier estimates for time to TB diagnosis, both crude and stratified by birth region; Kaplan-Meier estimates for the other covariates are located in Appendix C (Figures C1–C3). For all 4 variables, we identified statistically significant differences in the survival curves (p <0.01 for each). Table 2 and Figure 4 display the results of the Cox regression analysis: persons from Middle Africa had the highest hazard rate for developing TB disease, compared with persons from Western Europe, after adjusting for age at TB disease diagnosis, sex, and birth year (aHR = 7.02; 95% CI: 6.53–7.56); in other words, the chance of TB disease diagnosis is 7 times higher for non–US-born persons from Middle Africa than from Western Europe at any point during the study period, given that they did not previously have TB disease. Persons from all other regions had significantly higher adjusted hazard rates than persons from Western Europe at p ≤0.05, except for persons from Northern America (i.e., Canada). The complete results of the Cox regression analysis, including aHRs for age at TB disease diagnosis, sex, and birth year, are available in Table C1 (Appendix C). Finally, after performing our sensitivity analyses, we observed that increasing the exclusion window from ≤3 months to ≤6 months and excluding children aged <5 years changed the aHRs for all persons by <10%, regardless of birth region (Table C2, Appendix C).

**Figure 2.**
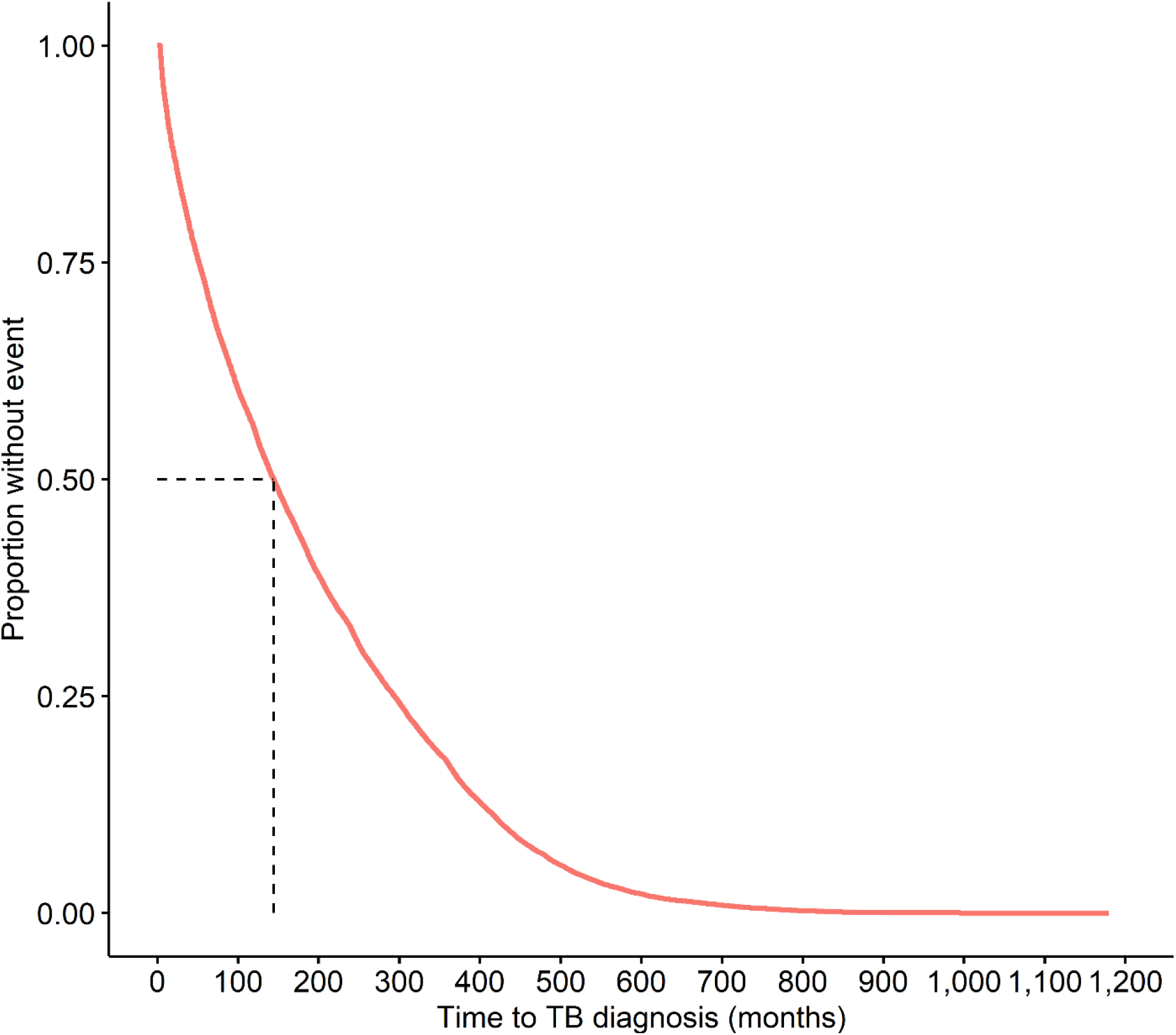
Kaplan-Meier estimates for time to tuberculosis (TB) disease diagnosis not attributed to recent transmission for non–US-born persons — United States, 2011–2018.

**Figure 3.**
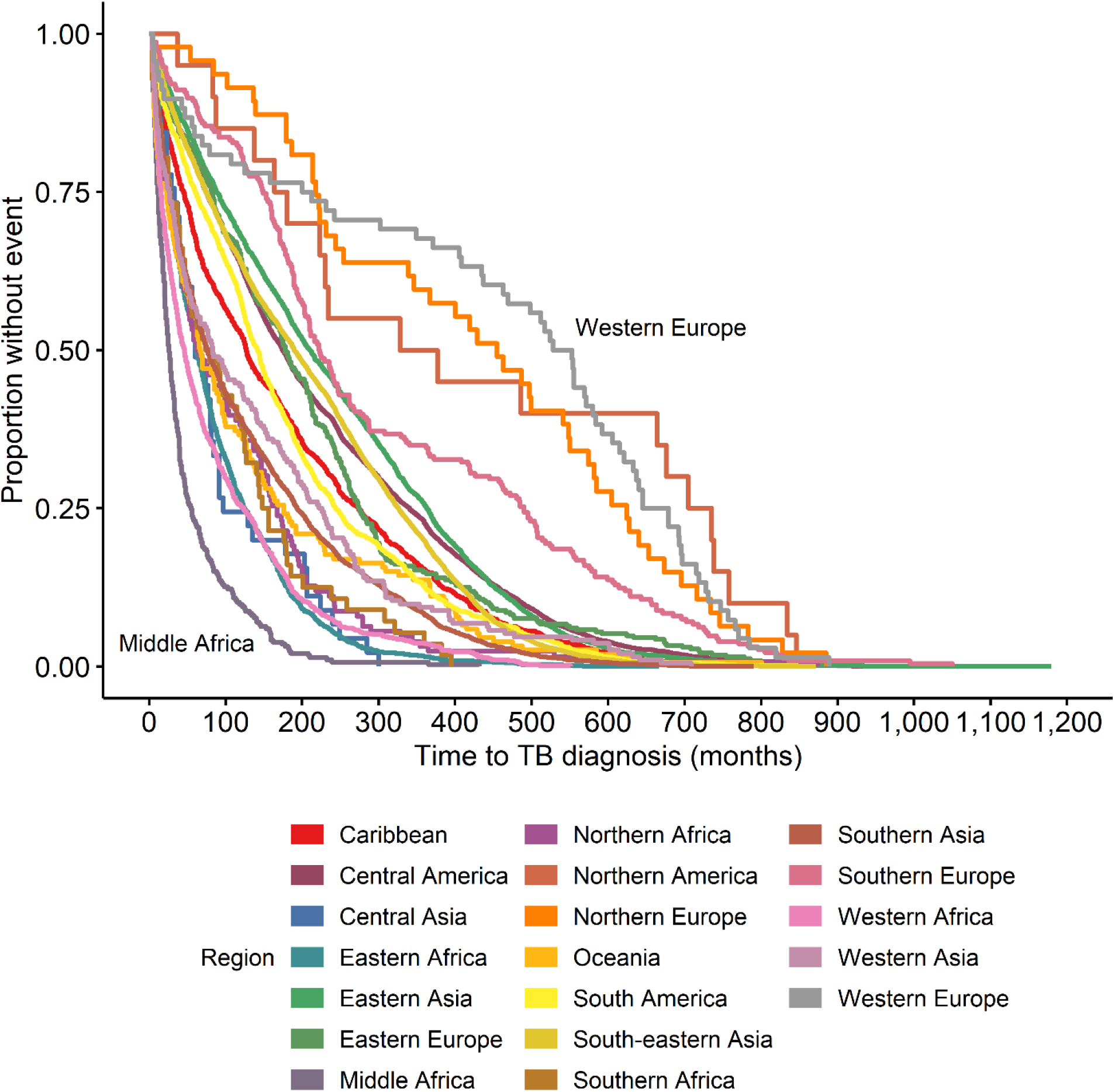
Kaplan-Meier estimates for time to tuberculosis (TB) disease diagnosis not attributed to recent transmission for non–U.S.-born persons stratified by region of birth (see Appendix A for list of countries) — United States, 2011–2018.

**Figure 4.**
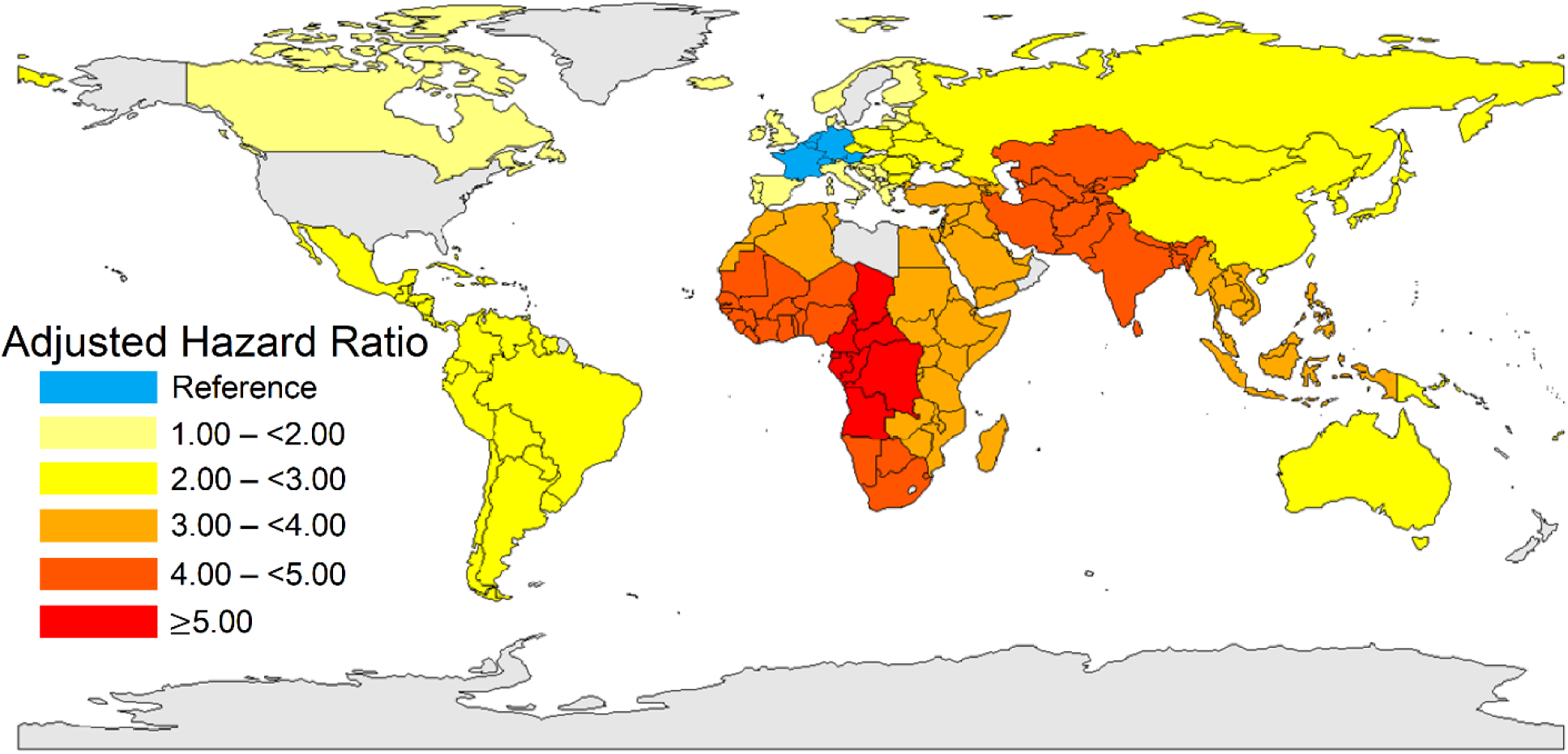
Map of adjusted hazard ratios for time to tuberculosis (TB) disease diagnosis not attributed to recent transmission for non–US-born persons according to region of birth (see Appendix A for list of countries) — United States, 2011–2018.

## Discussion

On the basis of our analysis of NTSS data, the length of time for non–US-born persons to receive a TB diagnosis varied according to birth region. The highest aHRs for TB disease diagnosis were concentrated among persons originating from Africa. In 2018, 5 of the 10 birth countries associated with the highest TB disease rates in the United States were African, and Africa had the highest TB disease rate globally (*1,18*). It is unclear if an actual causal association exists between higher overall TB disease rate in one’s birthplace and one’s likelihood of developing TB disease earlier. However, the World Health Organization’s African Region had the highest proportion of LTBI attributable to recent infection in 2014 (*19*), and persons are most likely to develop TB disease soon after TB infection (*5*). Therefore, shorter time to TB disease diagnosis might be associated with higher annual risk of TB infection. Another possibility is that US health care providers might have a higher suspicion for TB disease among certain populations, particularly persons from countries with high TB rates, a known risk factor for development of TB disease because of the higher risk for TB infection in those countries (*5*).

One other possibility is that these differences might reflect genetic differences between persons originating from different regions; certain host genetic factors have been associated with LTBI progression to TB disease (*20*). Although these potential etiologies are compelling, the differences observed in this study may likely be the consequence of overall poorer health status among persons from different regions, given lower levels of economic development and health care access; 33 of the 47 least-developed countries in the world are located in Africa (*21*), and Africa has the lowest overall healthy life expectancy (i.e., the number of years that a newborn is expected to live in good health) (*22*). Importantly, persons with poorer health status, particularly malnutrition (*23*), might be more susceptible to developing TB disease, compared with healthier persons.

The association between birth region and time to TB diagnosis may also be a consequence of differences in risk factors for progression to TB disease among different regions. Both HIV infection and diabetes mellitus are key risk factors for LTBI reactivation, with HIV infection representing the greatest risk factor for LTBI reactivation (*24*); worldwide, 9% of incident TB cases in 2018 were among persons living with HIV, and 15% of TB cases might be linked to diabetes mellitus (*18,25*). Additionally, the majority of high TB and HIV burden countries are in Africa (*16*), and the HIV diagnosis rates for African-born persons living in the United States are 6 times higher than the rate for the general US population (*26*). Given the possibility of TB and HIV coinfection or TB and diabetes mellitus comorbidity as plausible etiologies for the association between birth region and time to TB disease diagnosis, including HIV status or diabetes mellitus diagnosis as variables in our analysis might appear reasonable.

However, even though NTSS offers information on HIV status and diabetes mellitus diagnosis, it does not offer information on the date of testing or diagnosis. Therefore, to use these data, we would have to unjustifiably assume that a person’s HIV status or presence of diabetes mellitus were constant throughout the time from US entry to TB disease diagnosis date. This assumption would also be invalid because global HIV incidence was likely low before the 1980s (*27*), and before 2010, HIV infection could prevent non-U.S. citizens from entering the United States (*28*). In addition, including history of diabetes mellitus as a variable in this study would have resulted in an immortal time bias (*29*). For example, persons who received a diagnosis of diabetes mellitus during the study period (i.e., before receiving a diagnosis of TB disease) must have persisted long enough without a TB disease diagnosis to be diagnosed with diabetes mellitus; the period of time from the start of the study period to diagnosis of diabetes mellitus for these persons is known as *immortal time* because these persons might not have developed TB disease during that time interval. However, persons who have never received a diagnosis of diabetes mellitus might have received their TB diagnosis during this immortal time, resulting in a disadvantaged survival time despite not having a diabetes mellitus diagnosis. Therefore, we excluded HIV status and diabetes mellitus diagnosis as variables from this study.

Finally, all refugees entering the United States should undergo a domestic medical examination upon US entry. Given that Africa and Asia together account for the highest proportion of refugees coming to the United States since 2010 (*30*), their comparatively high aHRs in this study might be a consequence of TB disease diagnosis at the time of their domestic medical evaluation, especially since TB screening is a recommended component of that examination (*31*). However, the domestic medical examination is typically conducted 1–3 months after US entry (*32*), which is within our exclusion window for cases attributable to imported TB disease. In addition, all refugees are screened for TB before US entry, making it less likely that newly diagnosed TB would be detected following US entry.

## Limitations

Our study had several limitations. First, the characteristics of persons who immigrate to the United States may not represent persons who remain in their birth country. One study reported that birth country TB incidence was 5.4 times higher than the US TB incidence for persons born in those countries who immigrated to the United States (*33*); another study found that the prevalence by birth country of isoniazid resistance and multidrug-resistant TB in the United States better correlated with the prevalence by birth country seen in NTSS data than with the prevalence seen in the birth countries themselves (*34*). This suggests that non–US-born persons are not representative of the overall birth country population in terms of TB risk.

Additionally, NTSS does not report country of immediate origin, which can differ from birth country, nor does NTSS account for interceding travel outside the United States after initial US entry during which TB might have been transmitted. Finally, the health status of immigrants to the United States may be influenced by long-term US residence, causing it to diverge from expected health status on the basis of birth country.

Additionally, our study does not account for the number of non–US-born persons emigrating from a particular region nor how those numbers change over time. It also does not account for reason for immigration, such as refugee status, which may affect risk for TB infection. Likewise, it does not account for age at arrival or arrival year because adjusting for these variables with age at diagnosis would indirectly adjust for the outcome variable and cause the aHRs to converge to unity; adjusting for either of these variables would potentially account for time exposed to TB in a birth region, which may affect overall TB infection risk.

Furthermore, by using regions to categorize countries in this study, we lose the ability to detect differences among those countries. Also, although the plausible-source-case method employed by NTSS has high accuracy compared with field-based assessments of recent transmission that use epidemiologic investigation methods, it is not completely accurate and might misclassify certain cases (*3*). Finally, we were unable to assess the effect of HIV status and diabetes mellitus on time to TB disease diagnosis.

## Conclusion

Time to TB disease diagnosis among non–US-born persons varies by birth region, which represents an important prognostic indicator for LTBI progressing to TB disease. Targeted LTBI testing and treatment for persons entering the United States who were born in regions with high aHRs might advance progress toward TB elimination in the United States. Additional studies using data sources that include information on risk factors like HIV infection and diabetes mellitus would be helpful in determining the potential influence of these comorbid conditions on time to LTBI reactivation; such studies would also benefit from accounting for age at arrival and TB rates in birth countries where feasible. Similar studies in countries with health care systems comparable to the United States and diverse immigrant populations would also help determine if these results are reproducible and could shed light on the etiology underlying regional differences in time to TB disease diagnosis.

## Data Availability

The data contain information abstracted from the national tuberculosis case report form called the Report of Verified Case of Tuberculosis (RVCT) (OMB No. 0920-0026). These data have been reported voluntarily to CDC by state and local health departments, and are protected under the Assurance of Confidentiality (Sections 306 and 308(d) of the Public Health Service Act, 42 U.S.C. 242k and 242m(d)), which prevents disclosure of any information that could be used to directly or indirectly identify patients. For more information, see the CDC/ATSDR Policy on Releasing and Sharing Data (http://www.cdc.gov/maso/Policy/ReleasingData.pdf). A limited dataset is available at http://wonder.cdc.gov/TB.html. Researchers seeking additional data may apply to analyze National Tuberculosis Surveillance System data at CDC headquarters by contacting the Division of Tuberculosis Elimination.

## Acknowledgments

Andrew Hill; Carla A. Winston; Thomas R. Navin; Jonathan Wortham; Kristine M. Schmit; Sandy P. Althomsons; Noah Schwartz; Rebekah Stewart; Peter Cegielski; Andrew Vernon; National Center for HIV/AIDS, Viral Hepatitis, STD, and TB Prevention, CDC.

## Author Bio

Amish Talwar is an Epidemic Intelligence Service Officer in the Division of Tuberculosis Elimination at the Centers for Disease Control and Prevention in Atlanta. His primary research interests include infectious disease prevention and global health.

